# ChatGPT may free time needed by the interventional radiologist for administration / documentation: A study on the RSNA PICC line reporting template

**DOI:** 10.1101/2023.07.14.23292578

**Authors:** Jan F. Senge, Matthew T. McMurray, Fabian Haupt, Philipe S. Breiding, Claus Beisbart, Keivan Daneshvar, Alois Komarek, Gerd Nöldge, Frank Mosler, Wolfram A. Bosbach

## Abstract

**Motive:** Documentation and administration, unpleasant necessities, take a substantial part of the working time in the subspecialty of interventional radiology. With increasing future demand for clinical radiology predicted, time savings from use of text drafting technologies could be a valuable contribution towards our field.

**Method:** Three cases of peripherally inserted central catheter (PICC) line insertion were defined for the present study. The current version of ChatGPT was tasked with drafting reports, following the Radiological Society of North America (RSNA) template.

**Key results:** Score card evaluation by human radiologists indicates that time savings in documentation / administration can be expected without loss of quality from using ChatGPT. Further, automatically generated texts were not assessed to be clearly identifiable as AI-produced.

**Conclusions:** Patients, doctors, and hospital administrators would welcome a reduction of the time that interventional radiologists need for documentation and administration these days. If AI-tools as tested in the present study are brought into clinical application, questions about trust into those systems eg with regard to medical complications will have to be addressed.

## Introduction

In radiology, interventional radiology (IR) is the subspecialty which uses imaging for guiding minimally invasive surgical procedures. Imaging modalities applied today include fluoroscopy, ultrasound (US), computed tomography (CT), or magnetic resonance imaging (MRI) [1]. Our study investigated how IR could benefit from automated text drafting tools. We tested for the template of the Radiological Society of North America (RSNA) for peripherally inserted central catheter (PICC) lines [2] whether reports can be drafted by artificial intelligence (AI) based natural language processing models, ie ChatGPT [3].

Today, the interventional radiologist spends a substantial amount of time on administration / documentation work. As in other medical fields [4], [5], this activity is seen as an unpleasant necessity. It does not serve the immediate patient outcome. At the same time, demand for clinical radiology services is predicted to continue to grow in the future to a level that might not be able to be met by the workforce in its size today [6], [7]. This is why time savings through the use of AI text drafting would be a valuable and welcome contribution to the future of IR, from the viewpoint of patients, doctors, and hospital administrators alike.

Initial steps towards the computing technology required for AI in IR and elsewhere can be found in the work of Konrad Zuse [8]. The development of AI using digital computers was first proposed in eg [9], [10]. As other subfields of AI, natural language processing in combination with reinforcement learning, has recently seen some remarkable advances [3], [11]. This is a broader development, not limited to the tool applied in the present study [12]. Regarding the application in radiology and elsewhere, the strengths of properly trained language processing lie in the huge knowledge base that is made available [13], and in the ability to communicate in different styles of language [14]. So far, rather few studies on AI based language processing have been published in IR. One relevant study has demonstrated limitations concerning accuracy of recommendations for IR procedures [15]. This result is similar to what we found in a previous study about the handling of technical and medical information in report drafting for distal radius fracture [16], [17].

For evaluating the ability of ChatGPT to handle the RSNA PICC line template [2], we defined 3 distinct cases and iterated those for a parameter study (n = 5). Output texts were evaluated for content similarity and rated by 8 human radiologists. The main focus of the study was to determine if automation of text drafting seems feasible and will save time of the interventional radiologist.

## Method and Materials

The methodology of the presented study follows the concept of the previous work [16]: cases were defined within the framework of a current RSNA template. ChatGPT was tasked with report drafting. The output texts were evaluated for similarity by comparisons in python. The quality of output texts was assessed by human radiologists using a score card.

### RSNA template

The RSNA PICC insertion template can be found in [2]. Template items are listed in Table a. In the present study, three distinct cases were defined varying regarding eg anatomy, clinical information, and occurring complications. The impression had to be generated by the AI tool. “Patient ID”, and “Study ID” were added as parameters for the present study, in addition to the template items contained in [2].

**Table a:**
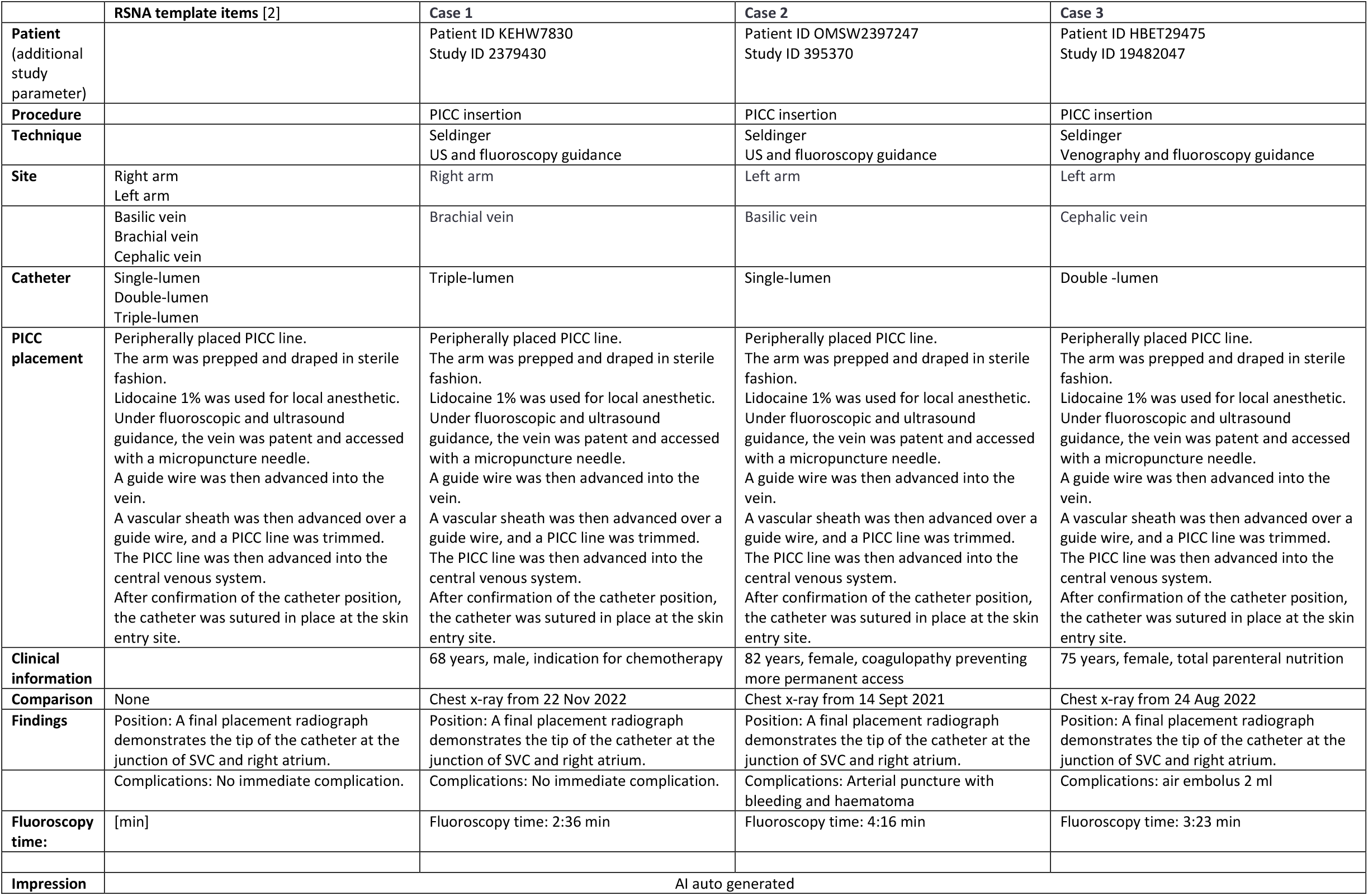
cases for defined parameter study.

### ChatGPT parameter study

The defined cases were given as command file to ChatGPT [3] on 04 May 2023 and iterated (n = 5), producing 15 output cases in total. The command was set to

> “*Write a radiology report which contains this exact information:*”.

No instruction was given on text structure, unlike before in [16]. The returned outputs were saved as txt-files. The previous study on distal radius fracture report drafting [16] relied on an earlier version of ChatGPT [18].

### Similarity analysis in Python

An analysis of similarity between text output files was performed following a method used before relying on bag of words in python: cosine similarity [0, 1] of vectors given by key word occurrence in command files defining the indicator vector space [16], [19].

### Score card assessment

Table b contains the structure of the score card given to radiologists participating in this study as raters. In total, 5 questions had to be answered for each of the 15 output texts. For this, raters had to grade on an ordinal scale [+2, +1, 0, -1, -2] how much they agree / disagree with the following statements:

**Table b:**
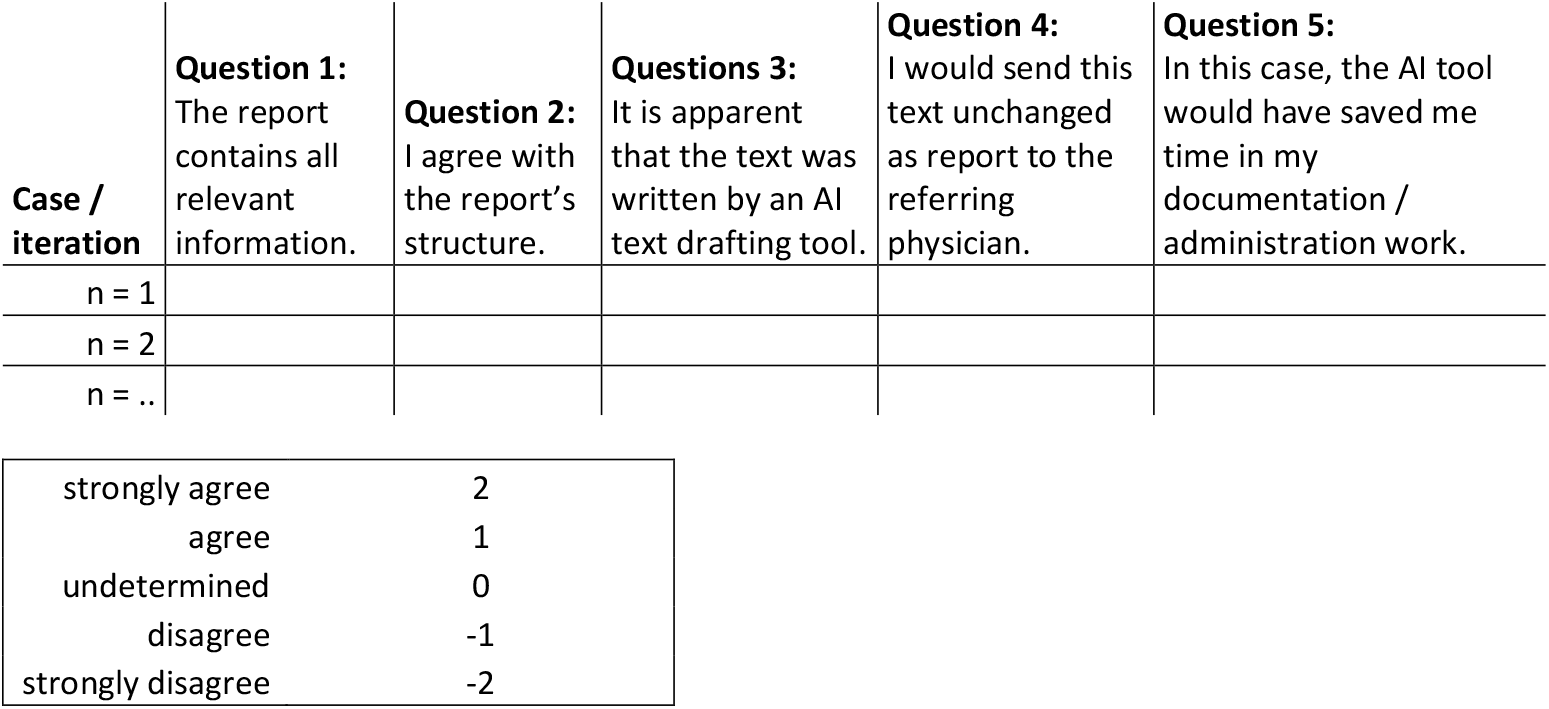
radiology scoring card.

1. The report contains all relevant information.
2. I agree with the report’s structure.
3. It is apparent that the text was written by an AI text drafting tool.
4. I would send this text unchanged as report to the referring physician.
5. In this case, the AI tool would have saved me time in my documentation / administration work.

Agreement regarding Questions 1, 2, 4, and 5 expresses a positive view on the ability of ChatGPT. As part of study’s design, Question 3 was deliberately worded to require disagreement from the rater for expressing a positive view on the ability of ChatGPT. Raters were blinded to the results of the other raters.

In total, 8 raters participated, 6 board certified radiologists, 2 residents. The total work experience averaged 22.5 years (min 6, max 49) for the board certified radiologists, with an average of 14.2 years within IR (min 1, max 34). Both residents were in their second year of residency training with 0.5 years in IR.

### Interrater agreement and reliability

For analysing the agreement and reliability between raters, a set of variables was calculated from the score card results, Table d. Each variable took values in the interval [-1, 1]. The approach followed the methodology used before in [16]. Three agreement measures were calculated: exact agreement, one-apart agreement, and weighted agreement with weights for ordinal scales defined in [20]. Chance-corrected interrater reliability variables for the present study included: Gwet’s AC1/AC2 (unweighted/weighted), the Brennan-Prediger coefficient, Conger’s kappa (generalization of Cohen’s kappa for multiple raters), Fleiss’ kappa, and Krippendorff’s Alpha. These coefficients can be defined via 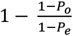, where of *P*_*O*_ and *P*_*e*_ are measures of observed and chance agreement, respectively. The different variables only differ in the definition of *P*_*O*_ and *P*_*e*_, for detailed formulas see [20]. Imbalance in the occurrences of certain (pairs of) scores in the overall crosstabulation matrix makes traditionally used kappa variables as well as Krippendorff’s Alpha prone to low reliability values. This paradoxon is further explained in [20]. Gwet’s AC and the Brennan-Prediger coefficient are less influenced by this imbalance effect. Computations were made using the package provided in [21].

## Results

### Sample output text

Table c contains the output example for case 1, iteration 1, defined in Table a. It can be seen that in principle ChatGPT can draft a PICC line report following the required input from the command file. Throughout the present study, output text structure varied compared to the example of Table c. ChatGPT repeatedly changed the contained section headings. A variation of output text structure was not seen before in [16] where text structure had been an explicit part of the command file.

**Table c:** output example case 1, iteration 1, generated by ChatGPT [3] for values defined in Table a for the RSNA PICC line template [2].

| ChatGPT output text, case 1, iteration 1: |
| --- |
| *Radiology Report:*<br>*Patient ID: KEHW7830*<br>*Study ID: 2379430*<br>*Procedure: Peripherally inserted central catheter (PICC) insertion under ultrasound and fluoroscopy guidance*<br>*Indication: Chemotherapy*<br>*Technique: The right arm was prepped and draped in sterile fashion. Lidocaine 1% was used for local anesthesia. Under fluoroscopic and ultrasound guidance, the brachial vein was accessed with a micropuncture needle. A guide wire was advanced into the vein, followed by a vascular sheath. A triple-lumen PICC line was trimmed and advanced into the central venous system. The catheter position was confirmed, and the catheter was sutured in place at the skin entry site.*<br>*Findings: A final placement radiograph demonstrates the tip of the catheter at the junction of SVC and right atrium.*<br>*Complications: No immediate complications were noted.*<br>*Fluoroscopy time: 2 minutes and 36 seconds.*<br>*Impression: Peripherally placed triple-lumen PICC line successfully inserted in the right arm brachial vein under fluoroscopic and ultrasound guidance. The catheter tip is positioned at the junction of SVC and right atrium. No immediate complications were noted.*<br>*Signed,*<br>*[Radiologist Name]* |

**Table d:** simple statistics of score card results, rater agreement, and interrater reliability.

| 1. simple statistics of score card results |  |  |  |  |  |  |
| --- | --- | --- | --- | --- | --- | --- |
| Question | Case | mode | median | range | mean | stdev |
| 1 | 1 | 2 | 2 | 1 | 1.8 | 0.40 |
|  | 2 | 2 | 2 | 4 | 1.43 | 0.92 |
|  | 3 | 2 | 2 | 4 | 1.35 | 1.06 |
| 2 | 1 | 2 | 2 | 4 | 0.90 | 1.55 |
|  | 2 | 2 | 2 | 4 | 1.23 | 1.23 |
|  | 3 | 2 | 2 | 4 | 1.63 | 1.07 |
| 3 | 1 | -2 | -2 | 3 | -1.48 | 0.84 |
|  | 2 | -2 | -1 | 4 | -1.13 | 1.21 |
|  | 3 | -2 | -1 | 4 | -0.95 | 1.30 |
| 4 | 1 | 1, 2 | 1 | 4 | 0.40 | 1.58 |
|  | 2 | 2 | 1 | 4 | 0.53 | 1.57 |
|  | 3 | 2 | 2 | 4 | 0.60 | 1.66 |
| 5 | 1 | 2 | 2 | 4 | 1.45 | 1.09 |
|  | 2 | 2 | 2 | 4 | 1.15 | 1.19 |
|  | 3 | 2 | 2 | 4 | 0.98 | 1.33 |
| 2. rater agreement in score card results |  |  |  |  |  |  |
|  | match | Question 1 | Question 2 | Question 3 | Question 4 | Question 5 |
|  | exact match | 0.49 | 0.49 | 0.35 | 0.25 | 0.39 |
|  | one-apart match | 0.88 | 0.73 | 0.74 | 0.49 | 0.64 |
|  | weighted match | 0.87 | 0.78 | 0.81 | 0.62 | 0.77 |
| 3. interrater reliability in score card results over all questions and cases |  |  |  |  |  |  |
| Coefficient name | value | weights | $P_o$ | $P_e$ | confidence interval | Benchmark: Landis-Koch |
| AC1 (identity)AC2 (weighted) | 0.27 | identity | 0.39 | 0.17 | 0.21 – 0.32 | Fair |
|  | 0.41 | weighted | 0.77 | 0.61 | 0.30 – 0.52 | Fair |
| Brennan-Prediger | 0.19 | weighted | 0.77 | 0.72 | 0.08 – 0.30 | Slight |
|  | 0.24 | identity | 0.39 | 0.20 | 0.19 – 0.29 | Slight |
| Conger's kappa | 0.33 | weighted | 0.77 | 0.66 | 0.23 – 0.44 | Fair |
|  | 0.13 | identity | 0.39 | 0.30 | 0.08 – 0.17 | Slight |
| Fleiss' kappa | 0.33 | weighted | 0.77 | 0.66 | 0.22 – 0.43 | Fair |
|  | 0.11 | identity | 0.39 | 0.32 | 0.06 – 0.16 | Slight |
| Krippendorff's Alpha | 0.33 | weighted | 0.77 | 0.66 | 0.22 – 0.44 | Fair |
|  | 0.11 | identity | 0.39 | 0.32 | 0.06 – 0.16 | Slight |

### Text similarity throughout the parameter study

Fig. 1 lists the headings of sections produced by ChatGPT and extracted from the 15 output files. With the exception of “Patient ID” and “Study ID”, no section heading appears in all 15 iterations. The average values included for Question 2 (I agree with the report’s structure) demonstrate some substantial variation between the 15 cases. Performance was particularly rated as poor whenever no section on complications was included. Within the set of 5 iterations for each of the three cases, score of Question 2 drops / increases whenever the section on complications is omitted / included by ChatGPT.

**Fig. 1:**
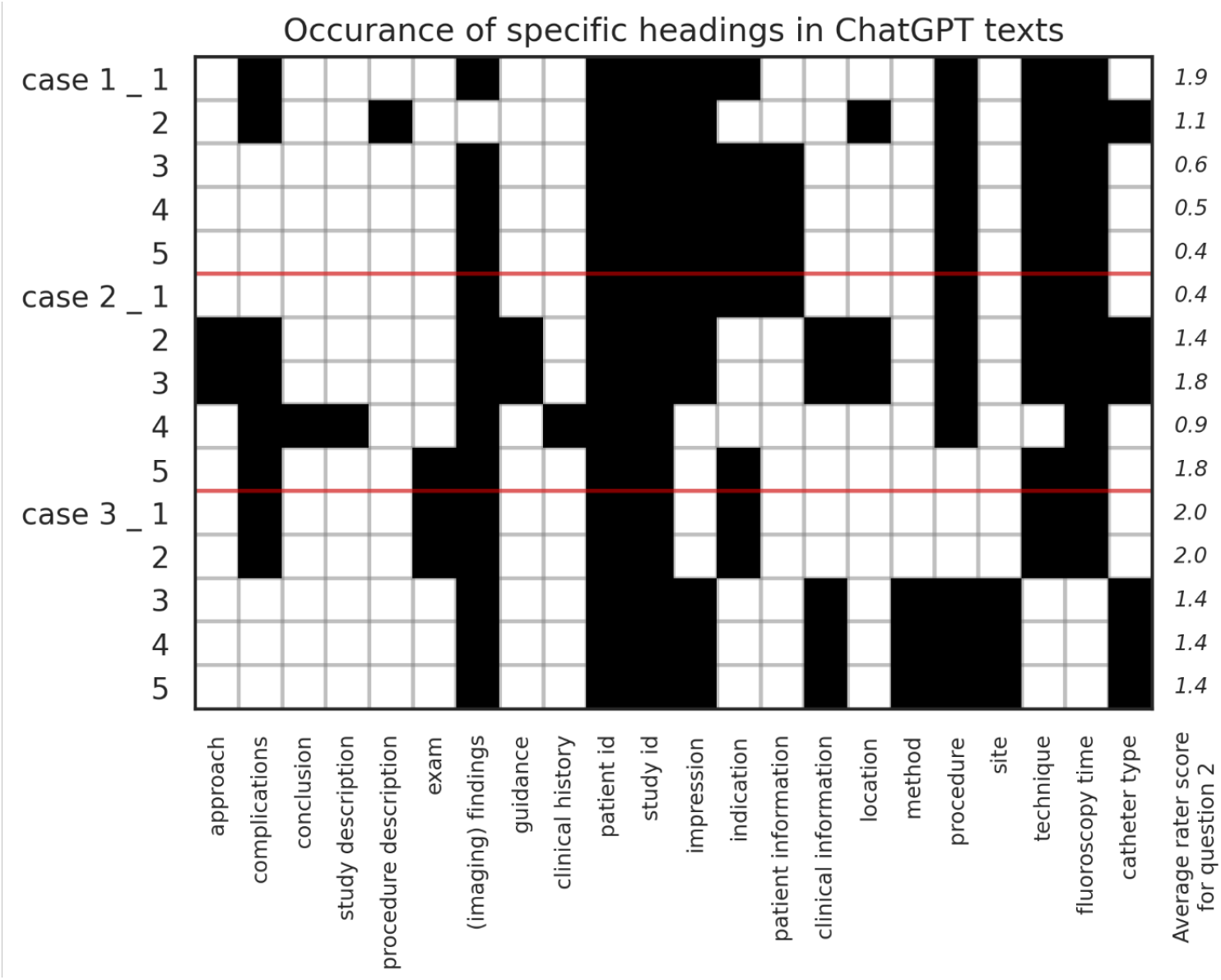
section headings extracted from the 15 output files, sorted alphabetically by second word in heading, together with average value from raters for Question 2: I agree with the report’s structure.

Fig. 2 provides a similarity comparison on a finer level and shows the cosine similarity calculated using bags of words. The comparison between the command files shows a [3, 3] matrix with the main diagonal taking the max value of 1, comparing the command files with themselves. Pairwise similarity between different command files lies between 0.75 and 0.80.

**Fig. 2:**
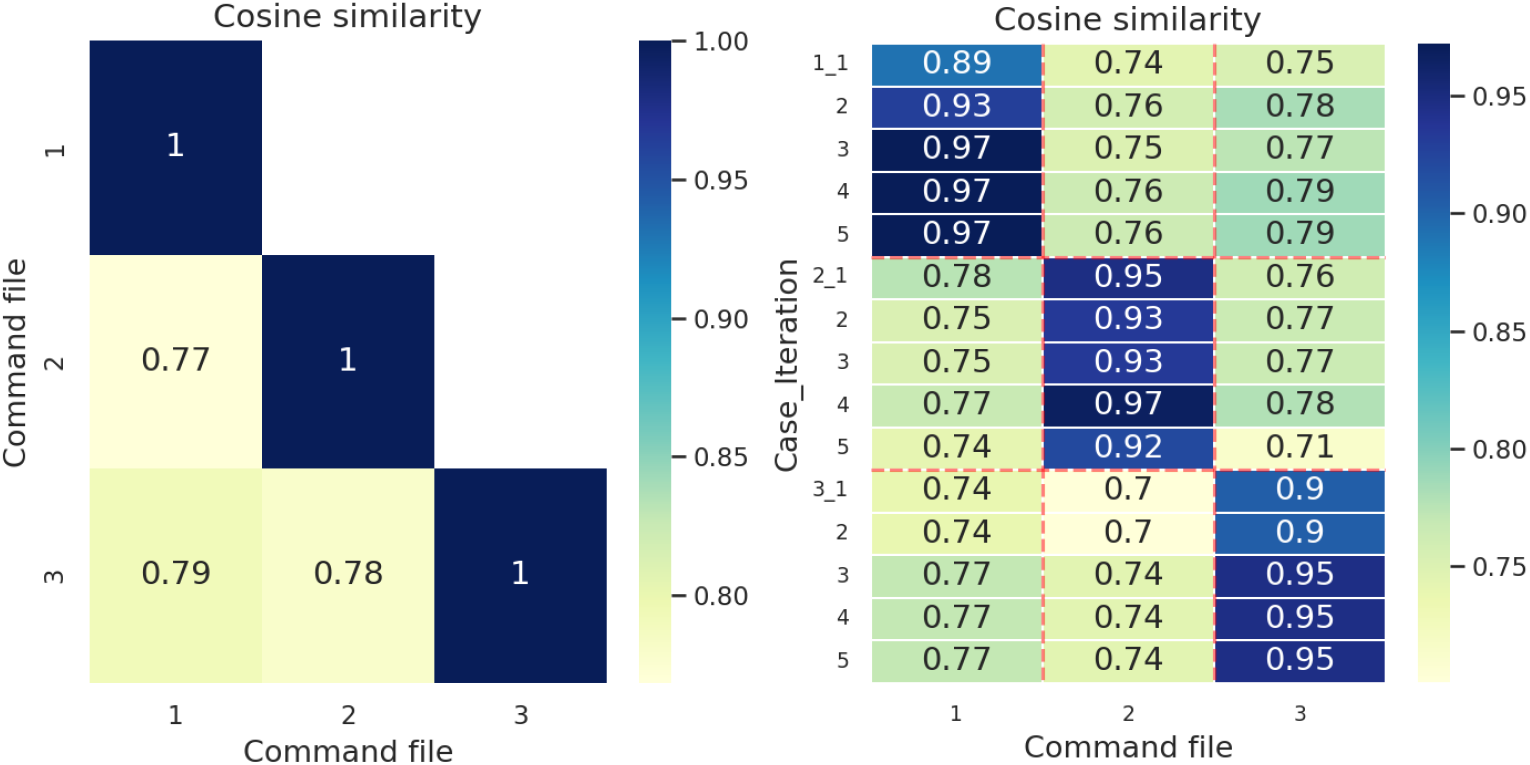
cosine similarity matrix between command files, and between command files and output files, computed by bag of words in Python.

The comparison between command files and output files is plotted as a [3, 15] grid. As before in [16], similarity exhibits plateaus of grid size [1, 5] along the main diagonal, resulting from comparison between each of the command files to the 5 corresponding output files. Outside these three plateaus, similarity drops substantially. This pattern was seen before in the previous study. It demonstrates again that ChatGPT has the ability to adjust its output to minor changes in the command file. Remarkably, the similarity of the ChatGPT output from one case to a command file of a different case is on average not much lower than the similarity between the respective command files. Accordingly, not much similarity is lost when we move from a command file to the output. While the [1, 5] similarity plateaus were highly homogeneous in the previous study, now on-plateau similarity varies markedly between values from 0.89 to 0.97. This, equally as before the change in text structure in Fig. 1, is new compared to [16] and can be attributed to the omission of prescribed text structure in the command file in the present study.

### Output text quality in scorecard assessment

Fig. 3 plots the distribution of the rater responses per question. Fig. 4 shows the average rater response with one standard deviation as error bar. Table d contains in its first panel the mode, median, range, mean and standard deviation.

**Fig. 3:**
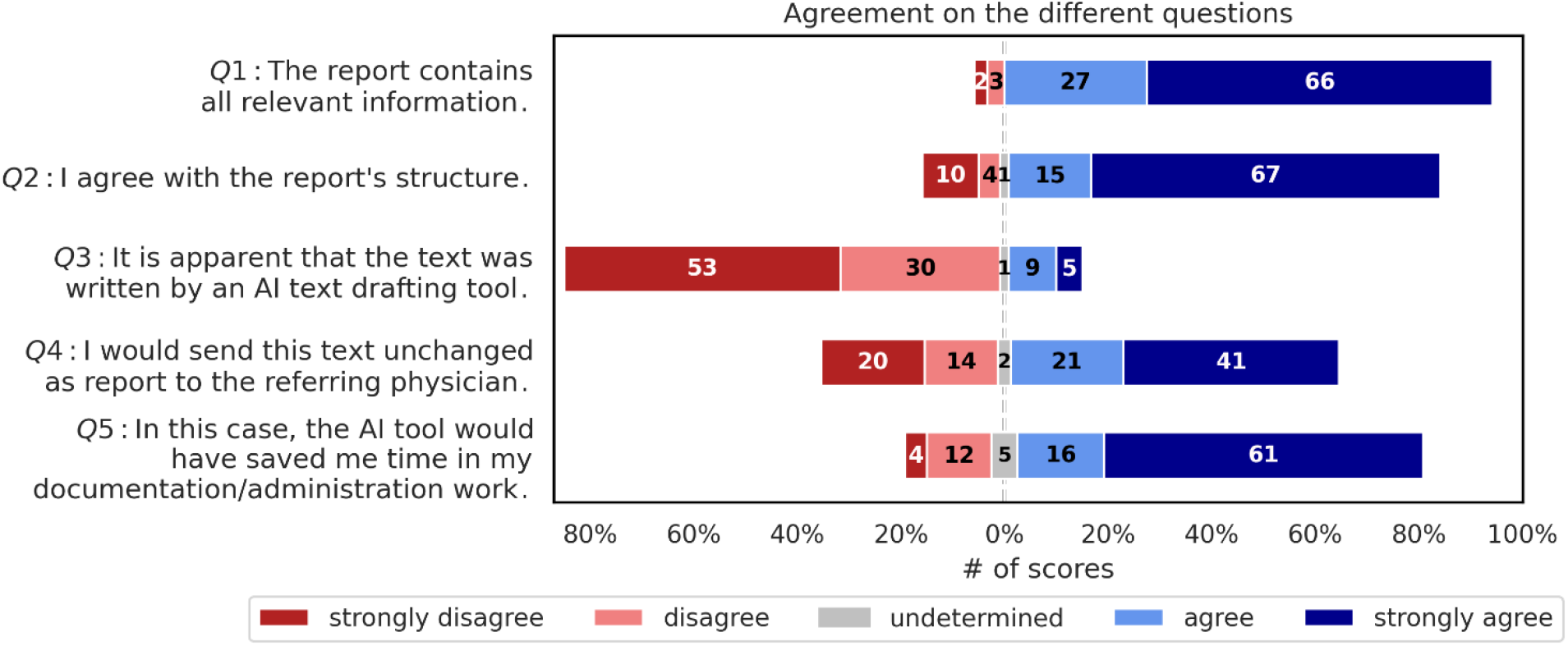
score card assessment with distribution of dis / agreement by raters per question.

**Fig. 4:**
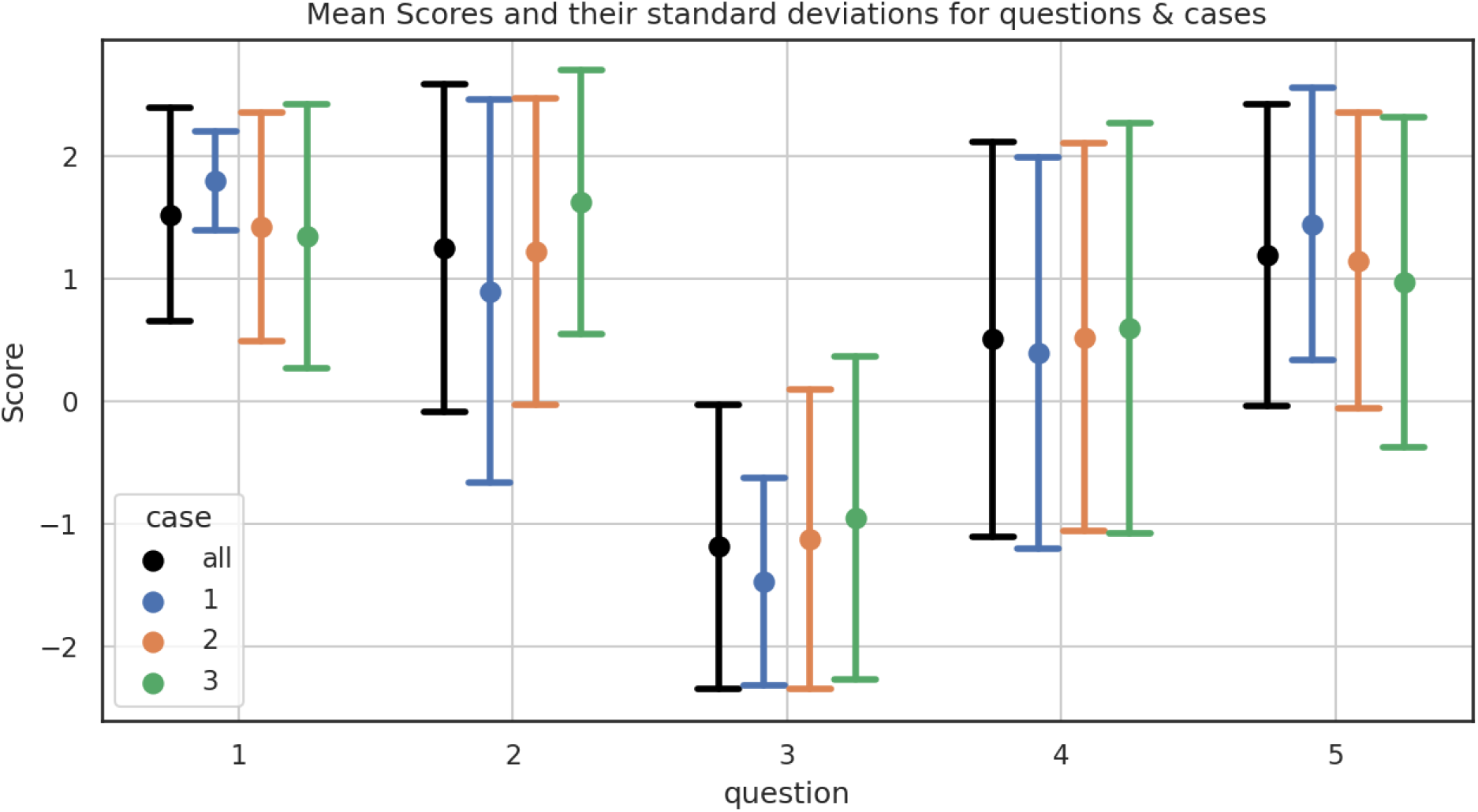
mean score with error bar of ± 1 standard deviation.

Overall, raters agreed with the statements offered in Questions 1, 2, 4, and 5; while disagreeing regarding Question 3. This can be interpreted as a clear positive statement about the quality of the AI generated PICC insertion reports.

Strong agreement was the most frequently given answer (strong disagreement in case of question 3 which was deliberately worded to require disagreement for a positive statement about the ChatGPT capabilities). By overall rater opinion, all relevant information was included (Question 1) in an agreeable text structure (Question 2). Raters overall disagreed with the statement that the output texts shown to them had been apparently written by an AI tool; accordingly, raters would not identify them as written by AI rather than by a human radiologist (Question 3). Question 4, whether the text draft could be sent out unchanged, saw a minor drop in mean agreement, as compared to the three previous questions. This indicates that raters would have considered editing the text draft manually before sending it. Question 5 received stronger agreement again, which affirms that, under the view of the participating human radiologists, AI-based automated text drafting will save time required in IR for administration / documentation.

Essential points raised by raters in their comments concerned text structure and handling of medical complications. Note that complications already influenced results of Question 2 and Fig. 1. Raters missed medical treatment suggestions which should have been included by ChatGPT in the PICC insertion report for the referring doctor by their opinion.

### Rater agreement and interrater reliability

An observation already made in [16] is confirmed by this study in Fig. 5 (standard deviation among raters, plotted over absolute rater mean; only negative means obtained under question 3 which required disagreement for a positive statement). Whenever texts are assessed to be of greater quality (abs(mean) ⟶ 2.0), variation between raters drops (standard deviation ⟶ 0.0). However, once quality is imperfect (abs (mean) ⟶ 0), there is an increasing variation between the raters’ expression of lack of agreement (standard deviation ⟶ 1.8). The point scatter in Fig. 5 can be interpolated linearly by regression analysis, R^2^ = 0.830. On the view of the authors, the pattern in Fig. 5 reflects real life situations in eg case presentations where proportion of disagreement between radiologists may increase with greater need for discussion. Section 2 of Table d contains the calculated rater agreement. Question 4 which received the lowest absolute mean also shows the lowest agreement between raters for all three agreement variables. This reflects the pattern observed before in Fig. 5 and discussed above. By definition, the agreement variables increase in most cases for wider defined range when calculated per question: exact match < one-apart match < weighted match.

**Fig. 5:**
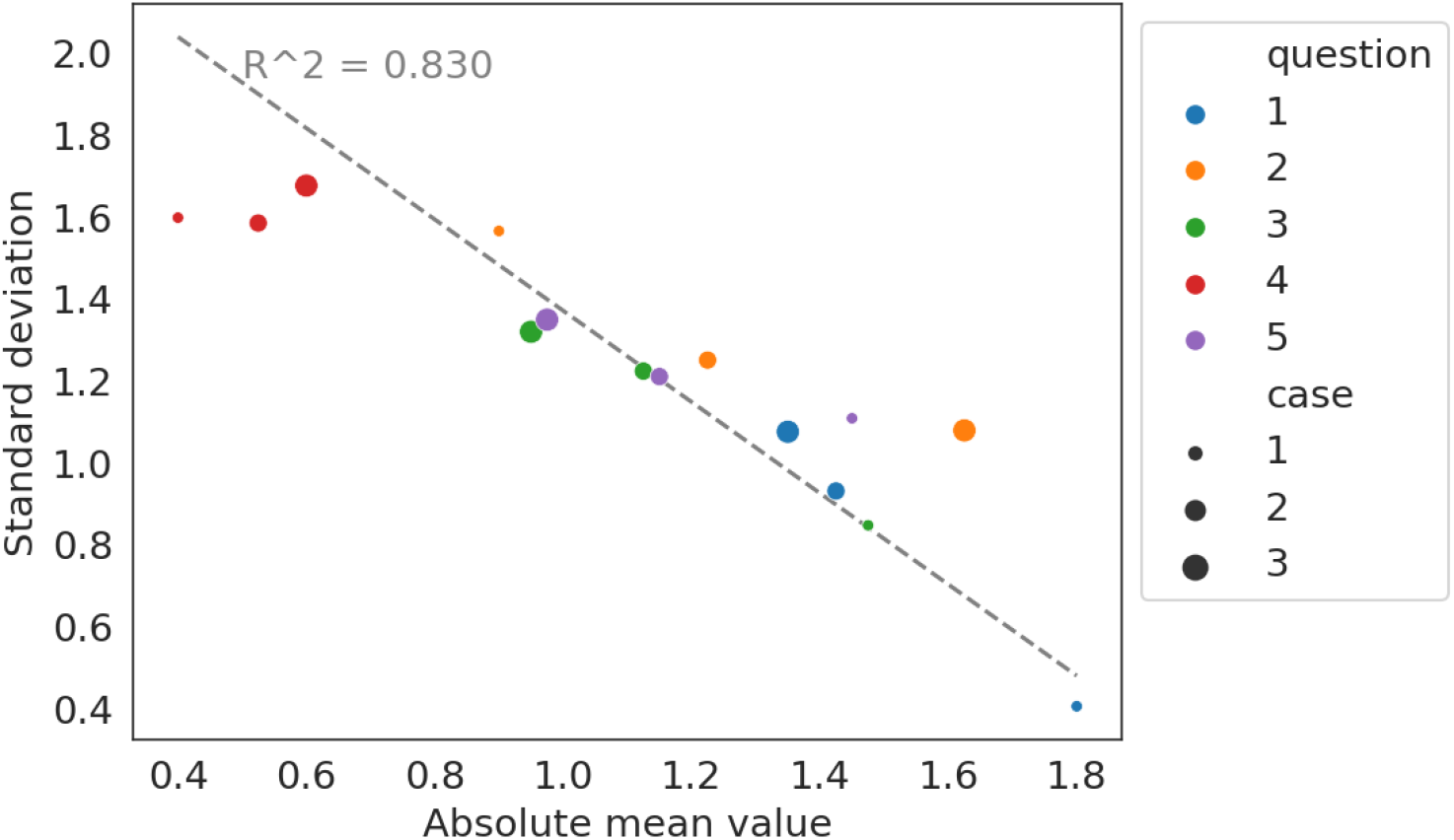
standard deviation plotted over absolute mean, aggregated per question per case, 15 data points.

Section 3 of Table d contains the calculated interrater reliability. Fair reliability was calculated for AC1 (unweighted / identity) and AC2 (weighted); as well as for weighted Conger’s kappa, Fleiss’ kappa, and Krippendorff’s Alpha. The remaining measures led to only slight reliability. This range of values is more consistent than what was obtained before in [16] for the evaluation of distal radius fracture reports. Most remarkable is the drop of AC1/2 from (identity: substantial, weighted: almost Perfect) in [16] to (identity: fair, weighted: fair) in the present study. Brennan-Prediger also saw a drop compared to the levels obtained in [16].

The interrater reliability between individual raters is shown as pairwise heatmap in Fig. 6 for weighted AC2. Raters are sorted for decreasing AC2 when calculating it for k raters. It can be seen that pairwise reliability reaches values of up to 0.95. Weighted AC2 decreases to 0.87 for the first four raters. When the remaining four raters of the total 8 are added, AC2 decreases to 0.41. Independently of the rates given, this finding too corresponds to real life experience according to which agreement between individual radiologists might well vary.

**Fig. 6:**
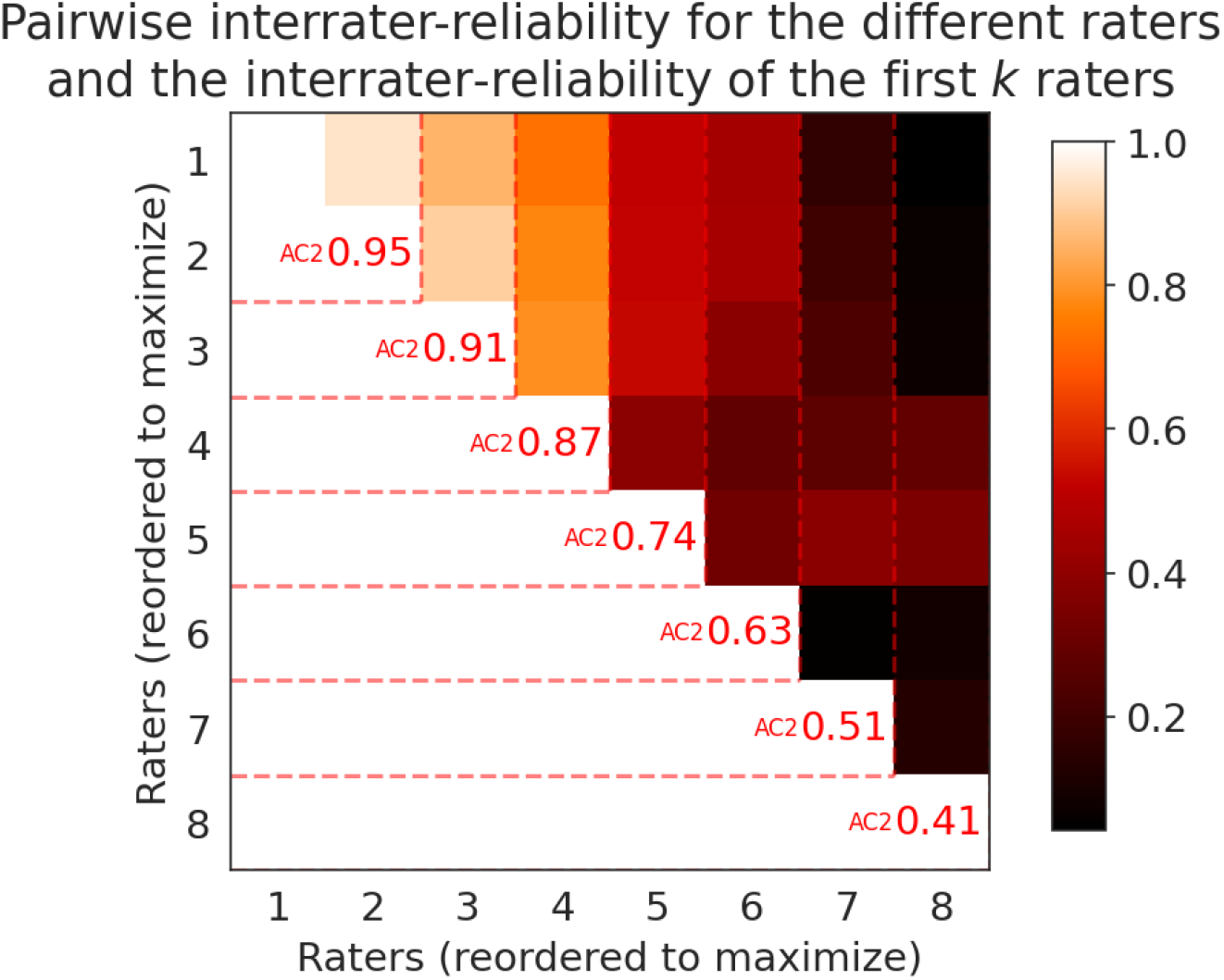
pairwise interrater reliability as heat map, as well was interrater reliability for group of the first k raters (red). The raters are sorted for descending magnitude of Gwet’s AC2 for greater group of raters.

Fig. 7 plots the reliability variables for each question. It demonstrates that AC2 and the Brennan-Prediger coefficient (except for Question 4) reached also in the present study greater values than the remaining variables, as before in [16]. The overall drop of AC2 and the Brennan-Prediger coefficient was effectively caused by question 4.

**Fig. 7:**
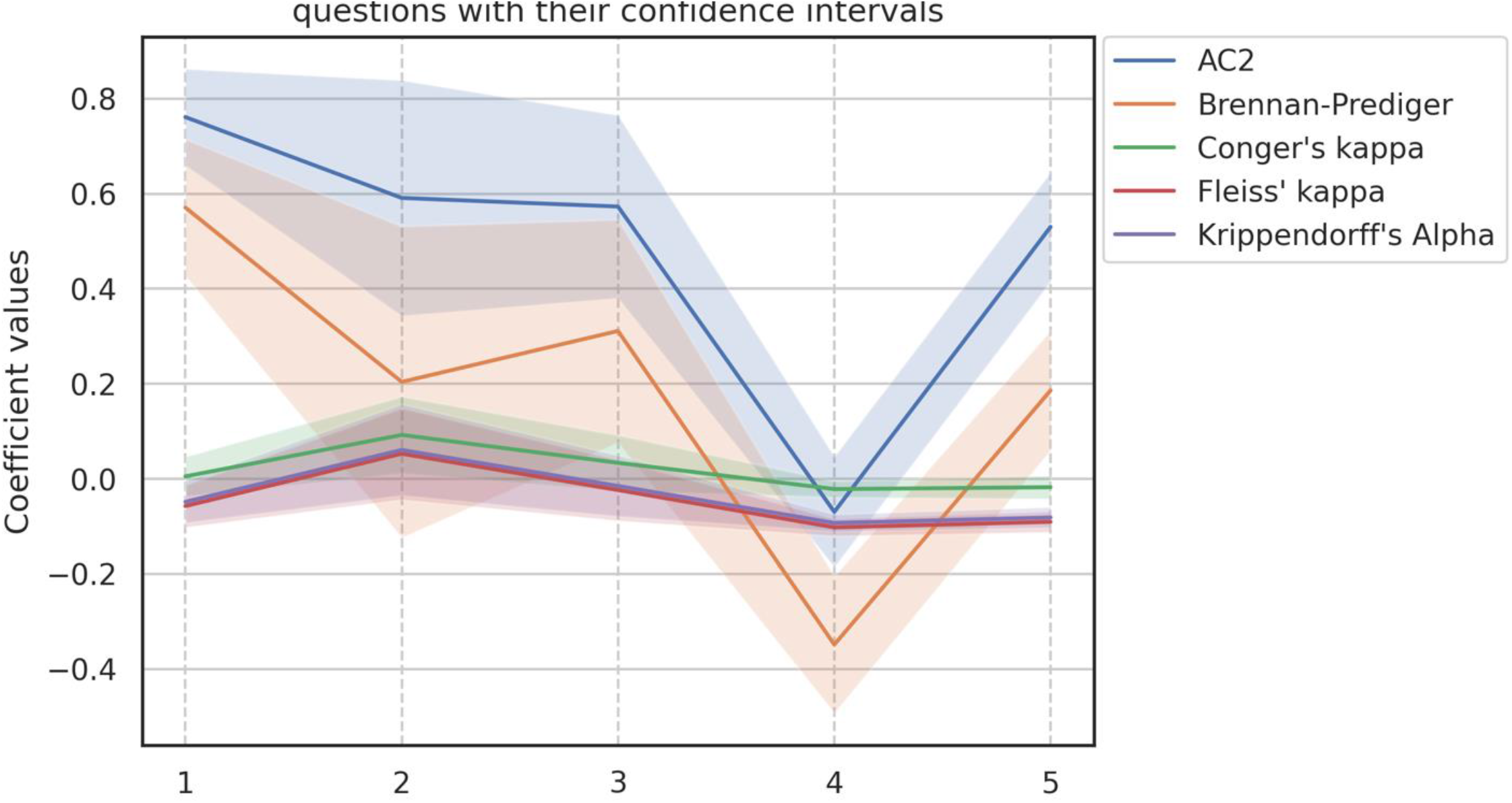
weighted Interrater reliability variables per question.

## Conclusions and future work

In the present study, we tested ChatGPT [3] for its ability to draft IR reports after PICC line insertion. Reports had to follow the current RSNA template [2] for three predefined study cases (Table a). Evaluation of the report drafts by human radiologists led to an overall positive assessment. One main result is: time savings in clinical administration / documentation of IR procedures can be expected from using ChatGPT (question 5). Future work will have to assess further the expectable magnitude of time savings when compared to today’s form of report writing which does typically not use AI generated drafts.

Overall, raters did not identify the output texts as written by an AI tool; this indicates that reports written by AI are for the raters indistinguishable from reports written by human radiologists (Question 3).

Due to the non-deterministic behaviour of ChatGPT, a parameter study was performed for each of the defined study cases (revisions n = 5). Unlike our previous study [16] in which text structure had been part of the input command, no required output text structure was given as part of the command file. As a result, the variation in text structure was stronger than in [16] (see Fig. 1). This drop in text similarity compared to [16] was also seen when calculating cosine similarity (see Fig. 2). Lack of reporting of complications as a separate report section by ChatGPT lowered scores on text structure (see Fig. 1).

In the set of scores received from the raters, a clear pattern could be identified (linear regression, R^2^= 0.83) that standard deviation increases for lesser absolute mean, Fig. 5. This pattern reflects real life situations where proportion of disagreement between radiologists may increase with greater need for discussion. Pairwise analysis of interrater reliability in Fig. 6 showed that also as in real life agreement between individual raters varied (max AC2 0.95, min AC2 0.41).

Mathematics in medical diagnostics is a wide field [22] with potentially many options for optimising healthcare and hospital operations, not limited to automation of clinical documentation [23], [24]. AI tools might well find their way into application and support the interventional radiologist in his administration / documentation tasks. Time savings, as can be expected from the results of the present study, would be an important improvement [4], [5]. Patients, doctors, and hospital administrators would agree on that.

Future work in this field will have to look deeper into ethical issues that may arise due to the application of ChatGPT in IR. One issue is whether professionals (radiologists, nurses etc.) trust AI-written reports. Also, patients may lose trust when they hear that reports are drafted using AI [25]. A second issue is how responsibility is shared between humans and AI [26]: Should humans stay in the loop? And who takes responsibility if something goes wrong? Finally, the privacy of patients is an issue because reinforcement learning uses input data to further train the model. Still, with continuing exposure of users and patients to AI tools and with steady improvements of technology and its ethical use, trust can be expected to grow.

## Data Availability

Online supplement: study raw data deposited under doi.org/10.5281/zenodo.8140755

## Acknowledgements and funding

The authors wish to thank for all the useful discussions leading to this manuscript.

## Ethics approval

not required

## Online supplement

study raw data deposited under doi.org/10.5281/zenodo.8140755

